# Mutations within the cGMP-binding domain of CNGA1 causing autosomal recessive retinitis pigmentosa in human and animal model

**DOI:** 10.1101/2022.08.10.22278420

**Authors:** Surabhi Kandaswamy, Lena Zobel, Bina John, Sathiyaveedu Thyagarajan Santhiya, Jacqueline Bogedein, Gerhard K.H. Przemeck, Valérie Gailus-Durner, Helmut Fuchs, Martin Biel, Martin Hrabĕ de Angelis, Jochen Graw, Stylianos Michalakis, Oana Veronica Amarie

## Abstract

Retinitis pigmentosa is a group of progressive inherited retinal dystrophies that may present clinically as part of a syndromic entity or as an isolated (nonsyndromic) manifestation. In a family suffering from retinitis pigmentosa, we identified a missense variation in *CNGA1* affecting the cyclic nucleotide binding domain (CNBD) and characterized a mouse model developed with mutated CNBD. A gene panel analysis comprising 105 known RP genes was used to analyze a family with autosomal-recessive retinitis pigmentosa (arRP) and revealed that *CNGA1* was affected. From sperm samples of ENU mutagenesis derived F_1_ mice, we re-derived a mutant with a *Cnga1* mutation. Homozygous mutant mice, developing retinal degeneration, were examined for morphological and functional consequences of the mutation. In the family, we identified a rare *CNGA1* variant (NM_001379270.1) c.1525G>A; (p.Gly509Arg), which co-segregated among the affected family members. Homozygous *Cnga1* mice harboring a (ENSMUST00000087213.12) c.1526A>G (p.Tyr509Cys) mutation showed progressive degeneration in the retinal photoreceptors from 8 weeks on. This study supports a role for *CNGA1* as a disease gene for arRP and provides new insights on the pathobiology of cGMP-binding domain mutations in *CNGA1*-RP.

## INTRODUCTION

Retinitis pigmentosa (RP) is a group of Inherited Retinal Degeneration/Dystrophies (IRD), with a global prevalence of 1 in 3000 – 7000 (1). RP is characterized by abnormalities in the photoreceptors (rods and cones) or the retinal pigment epithelium (RPE) with all types of inheritance patterns documented. RP can occur either as isolated or as syndrome with the involvement of other organs such as the associated hearing loss in USHER syndrome. About 90 genes are known until date to cause RP (Retnet database http://www.sph.uth.tmc.edu/retnet/). Most of the gene variants in RP are directly associated with the phototransduction cascade, such as *RHO* (rhodopsin), which are known to cause 25-30% of adRP. Phototransduction begins with the detection of light photons by rhodopsin, and this triggers several signaling steps that eventually convert the light signal into an electrical signal being transmitted to the brain. Key steps of this downstream signaling are mediated by proteins encoded by genes linked to RP. This list includes genes encoding for the subunits of rod phosphodiesterase (*PDE6A* and *PDE6B*) and rod cyclic nucleotide gated (CNG) channel (*CNGA1* and *CNGB1*). *CNGA1* encodes the A (or alpha) subunit of the rod CNG channel, which is a heterotetrameric channel complex formed by three CNGA1 and one CNGB1 subunits; its structure has been recently solved (2). The rod CNG channel along with CNGB1 forms a cyclic guanosine monophosphate (cGMP)-gated cation channel found in the rod photoreceptor outer segment plasma membrane (3). Each CNG channel subunit consist of six transmembrane domains, and both, the N-terminal and the C-terminal domain, are in the cytoplasm (2). While the A subunit is essential for the principle formation of a functional cGMP-gated channel, the B subunit is important for transport of the channel to the plasma membrane of the rod outer segment and confers specific properties to the channel complex such as rapid on-off kinetics and sensitivity to the pharmacological inhibitor L-cis-diltiazem (4,5).

In the present study we report a rare variant (c.1525G>A; p.Gly509Arg) of the *CNGA1* gene in a family suffering from arRP. Animal models for retinal degeneration usually provide insights into pathological mechanism of disease progression and assist in designing therapeutic strategies. We re-derived a *Cnga1* (c1526A>G; pTyr509Cys) mouse mutant from the ENU archive (6). This mutation falls within the same protein domain as the one observed in the human family. We report herein the retinal degeneration by a longitudinal morphological and physiological analysis.

## MATERIALS AND METHODS

### Clinical diagnosis and case recruitment

A male in his early 20s was registered with a complaint of night blindness at the retinal unit of the Eye Care Hospital. Detailed case history was recorded through a questionnaire designed for the study. Pedigree history revealed multiple affected family members including proband’s grandparent and two relatives. The study protocol was in accordance with the ethical guidelines of the 1975 Declaration of Helsinki. Written informed consent regarding publishing of their data and photographs, was obtained from all participants and study subjects were anonymized by specific internal codes. The study involving human subjects was approved by the Madras Medical College Institutional Ethical committee review board and all participants have signed a written informed consent when recruited (Approval no -29022013).

### Molecular analysis

To dissect the molecular pathology of the affected family, targeted retinal panel sequencing (TRPS) covering 105 genes involved in retinal dystrophies was performed at Medgenomics (Kochi, India); it included DNA library preparation, enrichment capture of exonic regions of the selected 105 genes (Supplementary table 1), cluster amplification, sample run on the Illumina Hiseq platform, and generation of raw data for further analysis. The data generated by TRPS were annotated and filtered based on the positive indels (above 35%), zygosity (homozygous/heterozygous), and analysis in the context of the disease (clinical documentation and phenotype) applying ACMG Criteria (7). The putative disease-causing variant was crosschecked among affected family members, among available unaffected family members and 120 unrelated control subjects of same ethnicity using custom-designed primers (Supplementary table 2) by direct sequencing.

### Structural comparison of CNGA1 wild-type protein versus mutant proteins (Human – CNGA1^Gly509Arg^; Mouse-CNGA1^Tyr509Cys^)

Structural models of CNGA1 proteins were generated using the RoseTTAfold deep learning algorithm (8) available at https://robetta.bakerlab.org/. The protein data bank (PDB) file 7RHH (9) was used as template structure. Sequences of human CNGA1 wild-type and respective mutant were obtained and based on protein isoform 2 (NP_000078.3). Sequences of mouse CNGA1 wild-type and respective mutant were based on NP_031749.2. Sequences were obtained from NCBI. Since the published sequence/structure is shorter than the known protein sequences, the wild-type and mutant sequences of both human and mouse CNGA1 were truncated accordingly. The generated 3D models were visualized using the UCSF Chimera software (https://www.cgl.ucsf.edu/chimera/).

### Generation of *Cnga1*^*Y509C*^ homozygous mice

ENU mutagenesis was performed as described previously (10). Briefly, ten-week-old C3HeB/FeJ male mice were injected intraperitoneal with ENU (three doses of 90 mg/kg in weekly intervals). First generation (F1) founder male mice were cryo-archived by their sperm and spleen-derived DNA samples. The DNA archive was screened for variants in the *Cnga1* gene affecting the cytoplasmic domain. The respective sperm sample was used to generate mice by *in vitro* fertilization and embryo transfer. The mutation was confirmed through PCR-based direct sequencing with custom-designed primers *Cnga1Ex9L1*: 5’-TGAGAGAGAAGTCCTGAGATACC-3’ and *Cnga1Ex9R1*: 5’-TGAGGTCATCTTTGGAGAGGC-3’ (Supplementary table 2). We established the *Cnga1* mutant line by repeated outcrossing with C57BL/6J mice to eliminate unwanted ENU mutations and the *Pde6b* mutation present in the C3HeB/FeJ genetic background (11). Mice were kept in pathogen-free quarters under a 12 h light/dark cycle and had *ad libitum* access to chow diet and water. All animal experiments were conducted in accordance with the German Law of Animal Protection and were approved by the government of upper Bavaria (Approval no - 55.2-1-54-2532-80-16).

### Spectral-domain optical coherence tomography

Optical coherence tomography was performed with an SD-OCT system (Spectralis®, HRA+OCT Heidelberg Engineering, Heidelberg, Germany) as described by Pawliczek et al. (12) and Schön et al. (13). Retinal sagittal sections were obtained along horizontal meridian, centered on the optic nerve. In addition, fundus BluePeak autofluorescence (BAF) and OCT angiography images (OCTA) were recorded. Mice were anaesthetized with ketamine (100 mg/kg)/xylazine (10 mg/kg) for the *in vivo* imaging and for the retinal function test. Mice were sacrificed by carbon dioxide.

### Electroretinography

Longitudinal ERG was performed in anesthetized animals (n = 8–14 per age group) after overnight dark adaptation using Celeris (Diagnosys LLC, Littleton, USA), as previously described by Wagner et al. (14). Briefly, pupils were dilated, and light guide electrodes were placed on both eyes centrally. Dark-adapted single flash intensity and flicker frequency series data were recorded. Stimuli of different light intensities were applied to the eyes, and the responses were recorded by the ERG device. Seven different stimuli ranging from 0.01 to 10 cd s m^-2^ were used for single flash measurements, which were based on the International Society for Clinical Electrophysiology of Vision (ISCEV) standardized protocol for clinical dark-adapted ERG recordings (15).

### Western blot

Protein lysates were obtained from retinas of 1 month old (PM1) and 6 months old (PM6) wild-type and *Cnga1*^*Y509C/Y509C*^ mice by disrupting the tissue with a mixer mill and extracting the proteins in RIPA lysis buffer (Merck, Darmstadt, Germany). Equal amounts of proteins were separated using 6-12 % SDS-PAGE, followed by Western blot analysis according to standard procedures. The following antibodies were used: rabbit anti-CNGA1 (1µg/mL), custom-made against the 19-mer Cys-RLTKVEKFLKPLIDTEFS-HN2, corresponding to 727–744 of human CNGA1 (NP_001136036.1); rabbit anti-CNGB1 1µg/mL, #4678 (16), anti-β-actin-peroxidase 1:25000 (A3854, Sigma-Aldrich, Saint Louis, USA), mouse anti-rabbit IgG-HRP 1:2000 (Sc-2357, Santa Cruz Biotechnologies, USA). Quantification was done using Image Lab Software Version 5.0 (Bio-Rad Laboratories, Munich, Germany).

### Histopathology

Eyes obtained from PM1, PM2 and PM4 controls and mutant mice were fixed 24 hrs in Davidson solution, dehydrated in 100% ethanol for 3 times (each for 15 min), embedded in Technovit® 8100 (Heraeus Kulzer, Wehrheim, Germany) and kept for polymerization for 6-10 hours at 4°C. Sagittal 2 µm sections through the middle of the eye ball were stained with basic fuchsine and methylene blue (BF&Me). Slides were scanned (NanoZoomer 2.0HT digital slide scanner, Hamamatsu, Japan) and taken images were processed with Adobe Illustrator image-processing program (17).

### Immunohistochemistry

Immunohistochemical staining was performed on sagittal cryo-sections of PM1, PM3, PM6, PM9 and PM12 retinas (13,16). We used the following primary antibodies: rabbit anti-CNGA1 (1:3000), custom-made against the peptide Cys-RLTKVEKFLKPLIDTEFS-HN2, corresponding to 727–744 of human CNGA1 (NP_001136036.1), rabbit anti-CNGB1 1:5000, #4678 (16),custom-made against the lectin anti-PNA FITC-conjugated 1:500 (#L7381, Sigma-Aldrich, Saint Louis, USA) and DAPI 1µg/mL. Laser scanning confocal micrographs were collected using a Leica SP8 confocal system (Leica, Wetzlar, Germany) equipped with the following lasers: 405, 448, 514, and 552 nm. Images were acquired as confocal z stacks using LAS X software V3.5.1.18803 (Leica). Maximum projection (merging of all z stacks) and background subtraction (value of 30) was performed using Fiji ImageJ V2.1.0/1.53c software (18).

### Real-Time quantitative Reverse Transcription PCR

Retinas were dissected and total RNA was isolated using the RNeasy Plus Mini Kit (Qiagen, Hilden, Germany). First-strand cDNA was synthesized from equal amounts of RNA with the RevertAid First Strand cDNA Synthesis Kit (Thermo Fisher Scientific, Waltham, USA). RT-qPCR was performed using the QuantStudio™ 5 Real-Time PCR System (Applied Biosystems, Taufkirchen, Germany). PowerUp SYBR Green Master Mix (Applied Biosystems) was used for quantification of amplified PCR products using specific primers for *Cnga1* (forward 5′-CTGTGAAGCTGGTCTGTTGG-3′; TAACTGCCGTCACTCAACAC-3′), 5′-TCTGAACAGGTGTCAGGATGTT-3′; reverse *Cngb1* 5′-(forward reverse 5′-CTGTTTCTGGCTGTGGTCCT-3′) 5′-TCGCCGATGCCCATTCTTATC-3′; 5′-GGCCCCAACTTCCATCAT CT-3′). and *mALAS* (forward reverse

### Statistics

Statistical analyses were performed using Prism 9 (GraphPad Software, San Diego, USA). Results are given as mean ± SEM or SD as indicated. Measured values of p ≤ 0.05 were considered significant. It is defined precisely as follows: *p ≤ 0.05, **p < 0.01, ***p < 0.001.

## RESULTS

### Clinical and molecular analysis of the affected family

The male in his 20s patient had bilateral progressive loss of night vision. Family history revealed autosomal recessive inheritance (Fig. 1A, full pedigree can be requested from the corresponding author). Fundus examination of the proband revealed pigmentary deposits in the periphery and the macular region, attenuated arterioles and waxy pallor disc (Fig. 1B). OCT examination revealed thinning of all retinal layers and a medium reflective lumpy lesion from the RPE suggesting lipofuscin deposits between the RPE and the photoreceptor layer. The surrounding black area denotes the subretinal fluid collection (Fig. 1C). This family was clinically diagnosed with early onset retinitis pigmentosa.

**Fig. 1.**
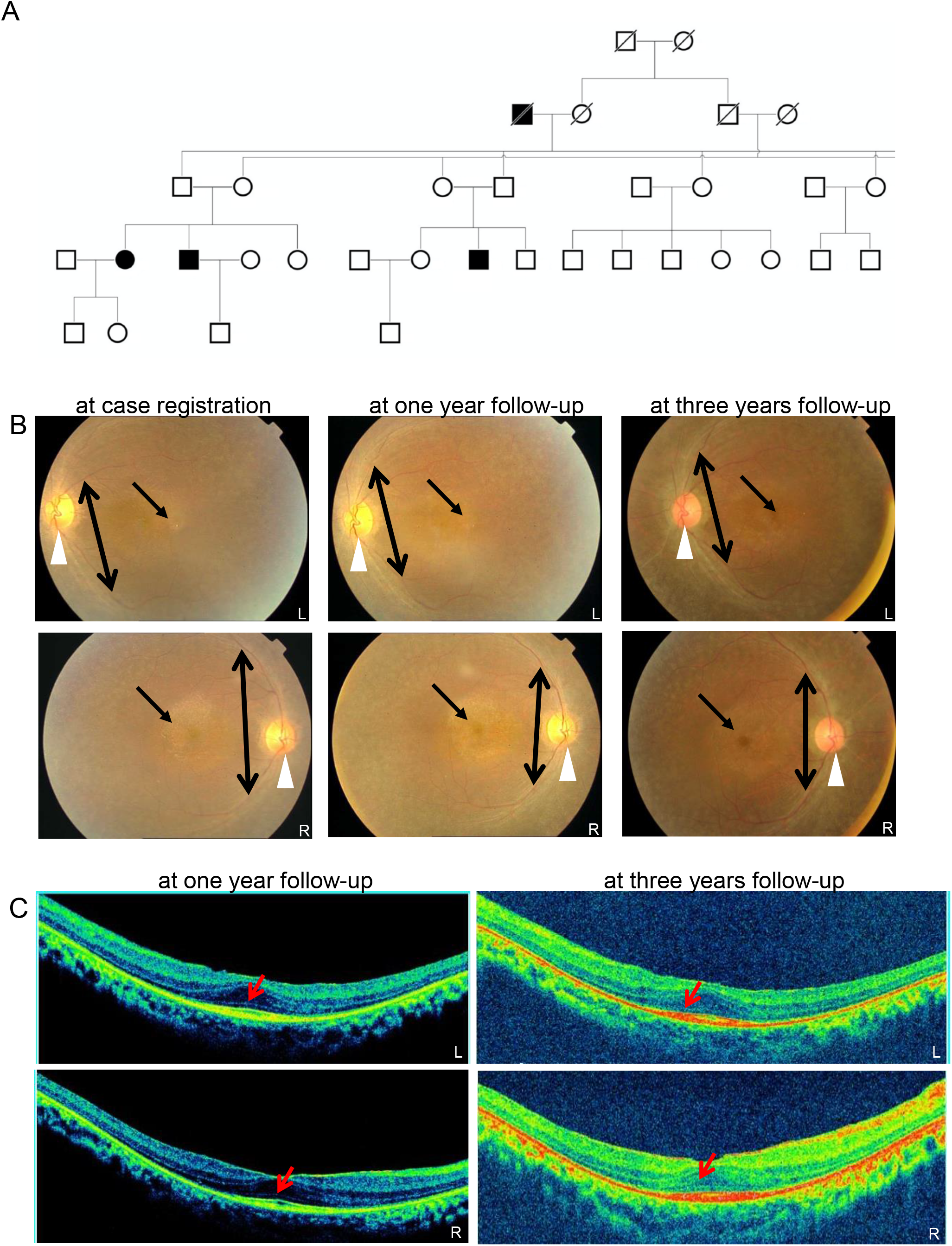
Pedigree and retinal imaging of the affected family: **A**. Partial five generation pedigree denoting the presence of ocular disease. Males and females, are represented by squares and circles, respectively. The symbols of affected family members are filled. **B**. Fundus images of the proband’s left (L) and right (R) eye taken at the time of case registration, on one year follow-up and again at three years follow-up. Waxy pallor disc (marked by white arrow head), pigmentary deposits (marked by one sided arrow) and attenuated arterioles (marked by double sided arrow). **C**. SD-OCT images of the proband’s retinal layers: left (L) and right eye (R) were taken on one year follow-up (left two panels) and three years follow-up (right two panels). Mild central hyper reflectivity suspicious of scars were marked with arrows. Thinning of the retinal layers was observed.

A rare homozygous variant in the gene *CNGA1*, (c.1525G>A; p.Gly509Arg was documented by TRPS (Fig. 2A). In affected family members, PCR based direct sequencing was resorted because no restriction site was available. The variant co-segregate amongst affected family members of proband, and two of his relatives, while the unaffected family members (proband’s parents, sibling, and the rest of the relatives were heterozygous carriers (Supplementary-Fig1)). This variant co-segregates amongst affected family members and is absent in 120 ethnically matched controls. The same variant ((NM_000087.3) c.1537 G > A; p.Gly513Arg) was earlier reported in association with another disease-causing variant as compound heterozygous condition underlying autosomal recessive retinitis pigmentosa in a Chinese family (19). Furthermore, this is also reported in the population database at a very low frequency (gnomAD exomes -0.000008029 (2/249086); (gnomAD genomes - 0.00001972 (3/152130) and no homozygotes were observed - Supplementary table 3). Both, the nucleotide (PhyloP score-5.885 and PhastCons score-1) and amino acid, are highly conserved across species (considering 14 species up to *C. elegans*) (Fig.2B). *In-silico* tools (Supplementary table 3), as well as applying ACMG criteria, predicted and classified this variant as pathogenic. To better visualize the mutated amino acid in the channel context, we performed RosettaFold-based modelling of wild-type and mutant CNGA1 proteins. Although RosettaFold only models the protein, not the cGMP molecule, it revealed that the mutant Arg509 (which is positioned at 509 in NP_001366199.1, p.Gly509Arg) protrudes into the inner space of the cGMP binding pocket (in Fig.2 C: compare the models with the cGMP-bound cryo-electron microscopy structure). Given that Arg is a basic amino acid, and occupies more space than Gly, which is neutral and allows for less flexibility of the amino acid chain, this substitution is expected to alter the overall secondary structure of the cyclic nucleotide-binding domain (CNBD) and the cGMP binding pocket (Fig.2 C).

**Fig. 2.**
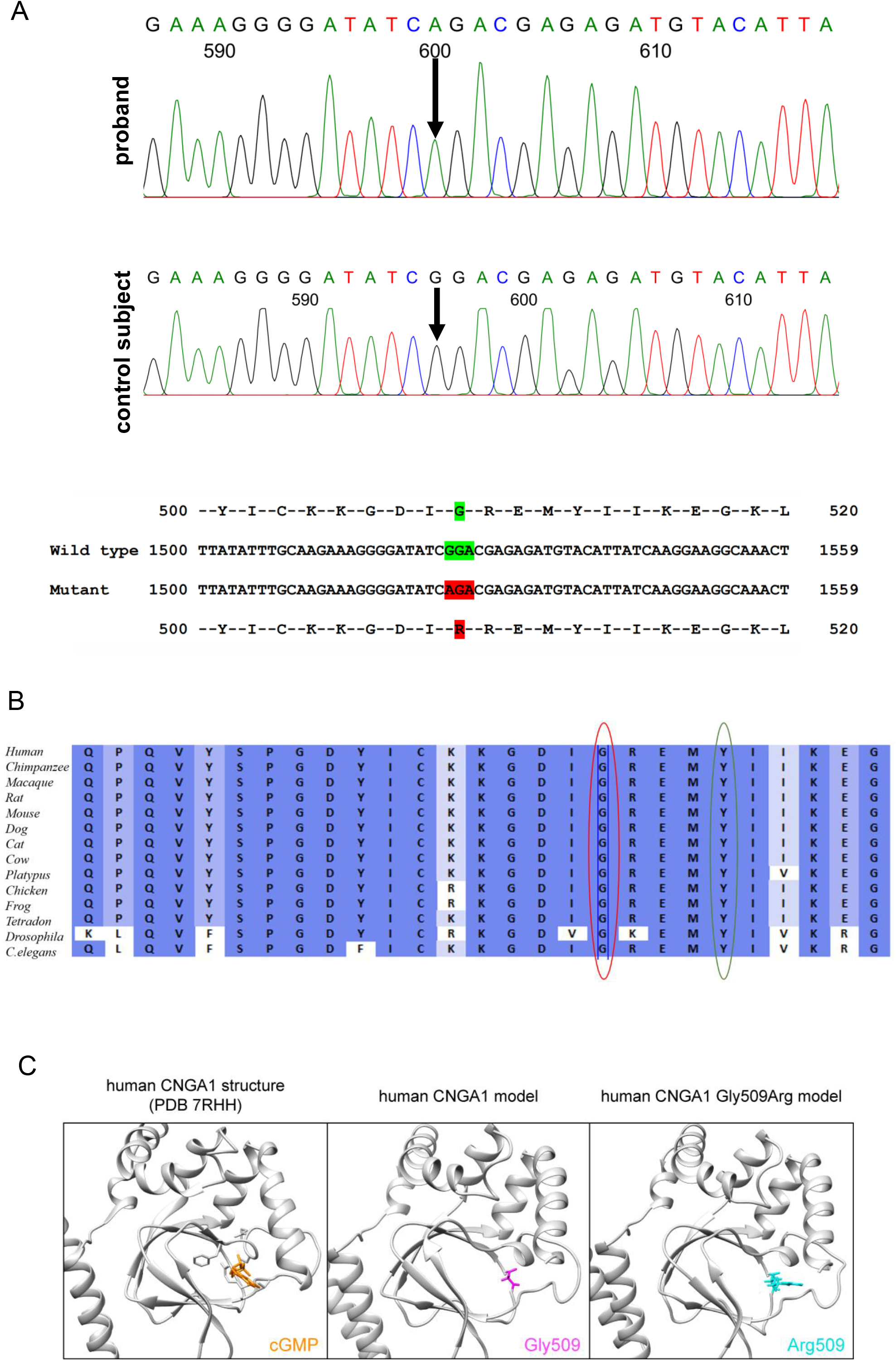
Identification of *CNGA1* mutation: **A**. Sequence chromatogram of the proband (top) and control subject from the regional population (middle) and *CNGA1* exon 10 (bottom), depicting the homozygous mutation (c.1525G>A; p.Gly509Arg); the mutant and wild-type peaks “A” and “G” are marked by arrows. The missense mutation (marked in red) is compared with the wild-type sequence (marked in green) together with the translated protein sequences. **B**. Amino acid sequence alignment of human CNGA1 and orthologues from other species, depicting a high conservation of p.G509 (encircled in red). Divergent amino acid residues are shaded in white color background. **C**. Structural comparison of wild-type and mutant human CNGA1 (backbone is shown in grey). The amino acid of interest in the wild-type structure (Gly513, magenta) and mutant structure (Arg513, cyan) (which is position at 509 in the MANE transcript encoded protein (NP_001366199.1) are shown as atoms. As reference structure the CNGA1 subunit of the human PDB 7RHH (9) is shown with bound cGMP (orange, shown as atom) and the residues R561, T562, A563, F544, E546, I547, S548 (grey, shown as atoms) are responsible for cGMP binding. Models were generated using the RoseTTAfold deep learning algorithm (8) available at https://robetta.bakerlab.org/. The generated 3D models were visualized using the UCSF Chimera software (https://www.cgl.ucsf.edu/chimera/).

### Generation of *Cnga1* mutant mice

From the DNA archive of ENU-induced mutant mice, we identified the missense mutation c.1526 A>G (Fig.3 A, B) in the *Cnga1* gene, leading to a substitution of Tyr509 by Cys. Mouse Tyr509 corresponds to Tyr513 in the human CNGA1 protein and thus affects a residue only 4 amino acids away from the mutant Arg509 in the human patient. Polyphen-2 predicted a probably damaging effect of this mutation (score 1.000), because this Tyr509 residue is highly conserved across a variety of species. Again, we performed RosettaFold-based modelling to visualize the wild-type and mutant CNGA1 proteins and again observed a slightly changed CNBD and cGMP binding pocket structure (Fig. 3 C).

**Fig. 3.**
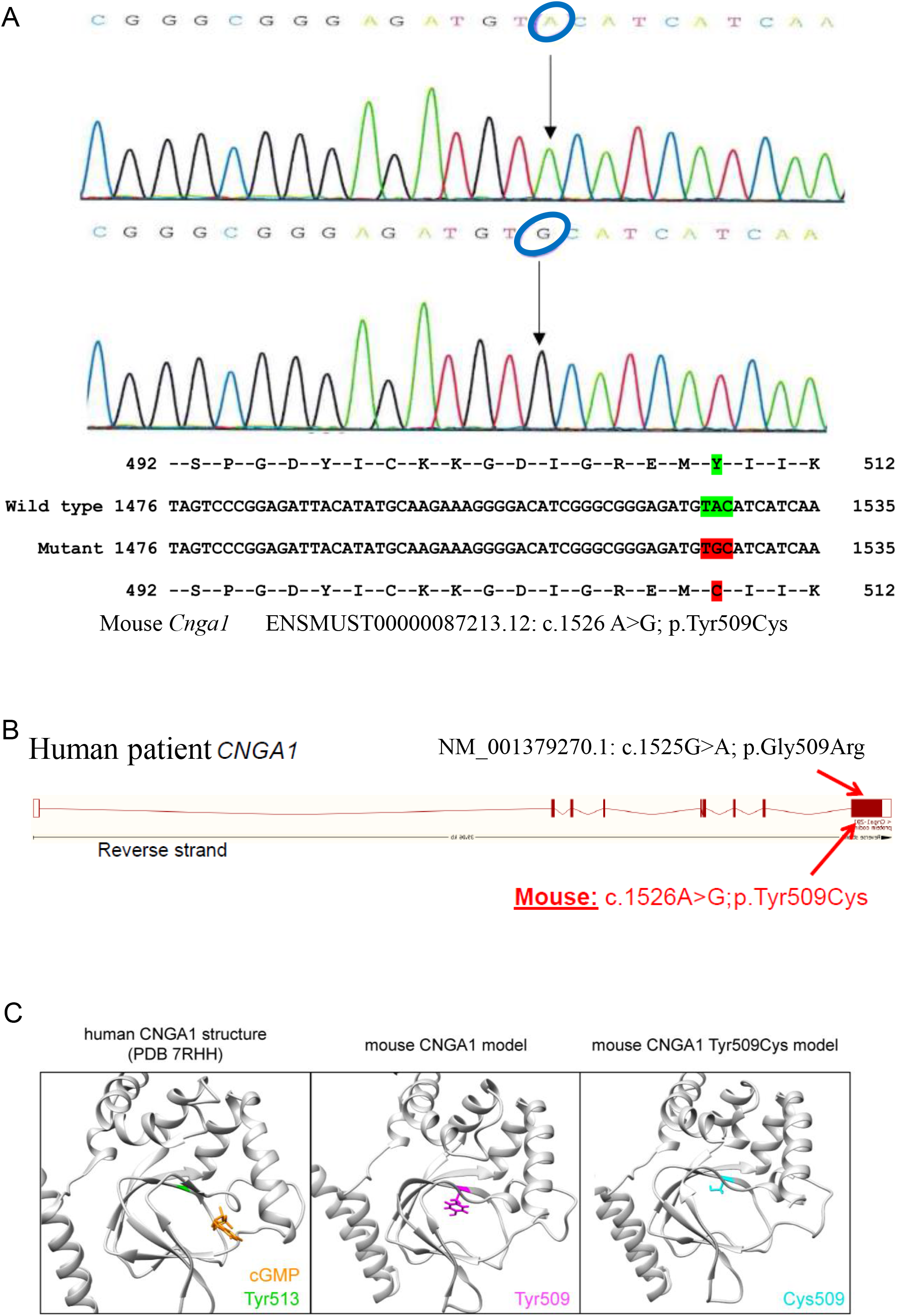
*Cnga1* mouse model: **A**. DNA sequencing shows the mutation in the new mouse model (c. 1526LJA>G; blue circles). The missense mutation (marked in red) is compared with the wild-type sequence (marked in green) together with the translated protein sequences of the mouse. **B**. Location of the human (p.Gly509Arg, top) and murine (p.Tyr509Cys, bottom) mutations in the cGMP-binding domain. **C**. Structural comparison of wild-type and mutant murine CNGA1 (backbone is shown in grey). The amino acid of interest in the wild-type structure (Tyr509, magenta) and mutant structure (Cys509, cyan) are shown as atoms. As reference structure, the CNGA1 subunit of the human PDB 7RHH [9] is shown with bound cGMP (orange, shown as atom) and the corresponding amino acid residue Tyr510 is highlighted (green). Models were generated using the RoseTTAfold deep learning algorithm (8) available at https://robetta.bakerlab.org/. The generated 3D models were visualized using the UCSF Chimera software (https://www.cgl.ucsf.edu/chimera/).

### Rod CNG channel expression in *Cnga1* mutant mice

We first analysed the effect of the mutation on the expression of the rod CNG channel homozygous *Cnga1* mutants. To determine the expression pattern of CNGA1, we immunolabeled retinal cross-sections of wild-type and *Cnga1* mutant mice for CNGA1 and CNGB1 proteins. In the wild-type retina, we observed a strong and specific immunofluorescence signal for both CNGA1 and CNGB1 in rod outer segments of one-month-old (PM1) mice (Fig. 4 A, D). In contrast, under the same imaging conditions, no CNGA1 signal was observed in the *Cnga1* mutant retina (Fig. 4 B). In six-month-old (PM6) mice, the CNGA1 immunosignal was also absent, in addition to a marked thinning of the outer nuclear layer (Fig. 4 C). Interestingly, the mutant retinas were also immunonegative for CNGB1 indicating degradation of the CNGB1 protein in the absence of CNGA1 (Fig. 4 E – F). Western blot analysis on retinal lysates detected the corresponding proteins in the wild type and confirmed the reduction of both CNG channel subunits in the mutant (Fig. 4 G – I, uncropped Western blots in Supplementary Figure 2). Analysis of the corresponding gene expression by qRT-PCR revealed similar levels of *Cnga1* mRNA transcript at 1 month of age and significantly decreased levels at PM6, when substantial ONL loss had occurred (Fig. 4 J). The levels of *Cngb1* mRNA were already slightly downregulated at PM1 and further decreased at PM6 (Fig. 4 K). These findings suggest that the *Cnga1* mutation does not impair principle gene expression and most likely exerts its negative effect at the protein level.

**Fig. 4.**
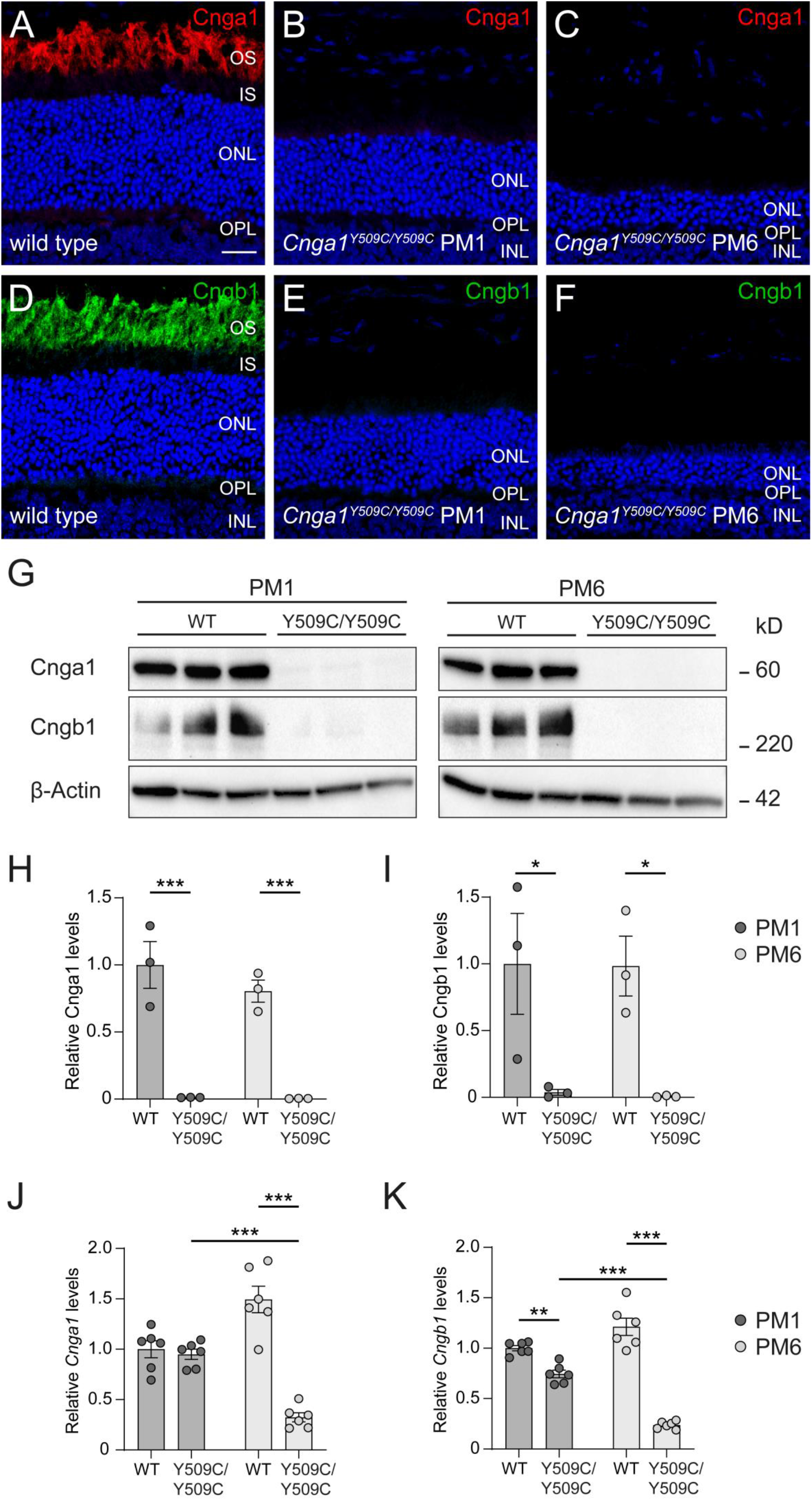
*Cnga1*^*Y509C/Y509C*^ mice are lacking CNGA1 protein. (**A-F**) Representative confocal images showing expression of CNGA1 (red) and CNGB1 protein (green) in retinal cross sections of wild-type (1 month postnatal (PM1); A, D), and *Cnga1*^*Y509C/Y509C*^ mouse retinas (B-C, E-F) at PM1 and PM6. *Cnga1*^*Y509C/Y509C*^ mice are lacking CNGA1 and CNGB1 already at PM1. Cell nuclei were stained with DAPI (blue). OS, outer segments; IS, inner segments; ONL, outer nuclear layer; OPL, outer plexiform layer; INL, inner nuclear layer. Scale bar marks 20 µm. (**G-I**) Western blot analysis of *Cnga1*^*Y509C/Y509C*^ mouse retinas at PM1 and PM6 using CNGA1- and CNGB1-antibodies. β-Actin was used as control. Western blot staining (G) and quantification of CNGA1 (H) and CNGB1 (I) expression confirm the findings of immunohistochemistry. (**J-K**) RT-qPCR of *Cnga1*^*Y509C/Y509C*^ mouse retinas at PM1 and PM6 with *Cnga1*-(J) and *Cngb1*-specific primers (K). Mutant mice at PM1 still express *Cnga1* and *Cngb1* transcript. This expression is reduced at PM6. N = 3 biological and technical replicates. Values are given as mean ± SEM (one-way ANOVA paired with Tukey’s post-hoc test; * * p≤0.01, * * * p≤0.001).

### Retinal function in *Cnga1* mice

Next, we assessed retinal function in *Cnga1* mutant mice by electroretinography (ERG). Mice were evaluated starting at postnatal week 3 (PW3), PM1, PM3, PM6, PM9 and PM12 to track functional changes over time. Scotopic and photopic ERG protocols were applied to evaluate rod- and cone-mediated light responses. Representative scotopic ERG traces and corresponding quantifications of the ERG a- and b-wave amplitudes are shown in Fig. 5. In wild-type mice, a- and b-wave amplitudes were clearly visible and showed an expected slight age-dependent decline over time (Fig. 5 B and E). In contrast, we could not detect any rod-derived a-wave in *Cnga1* mutant mice (Fig. 5 B, C and D), suggesting that mutant rods are incapable of generating light responses. The ERG b-wave was also absent after stimulation with rod-specific low luminance (0.01 or 0.03 cd.s/m^2^) and decreased strongly at higher luminance (Fig. 5 E, F and G). From PM9 onwards, *Cnga1* mutant mice no longer respond to light stimuli, since no ERG response was detectable even at the highest luminance (10 cd.s/m^2^) (Fig. 5 A, E and G). In summary, ERG analysis indicates a lack of rod photoreceptor-driven responses in *Cnga1* mutant mice as early as PW3 and reveals a secondary, slowly progressive loss of cone-mediated light responses, leading to complete blindness after PM9.

**Fig. 5.**
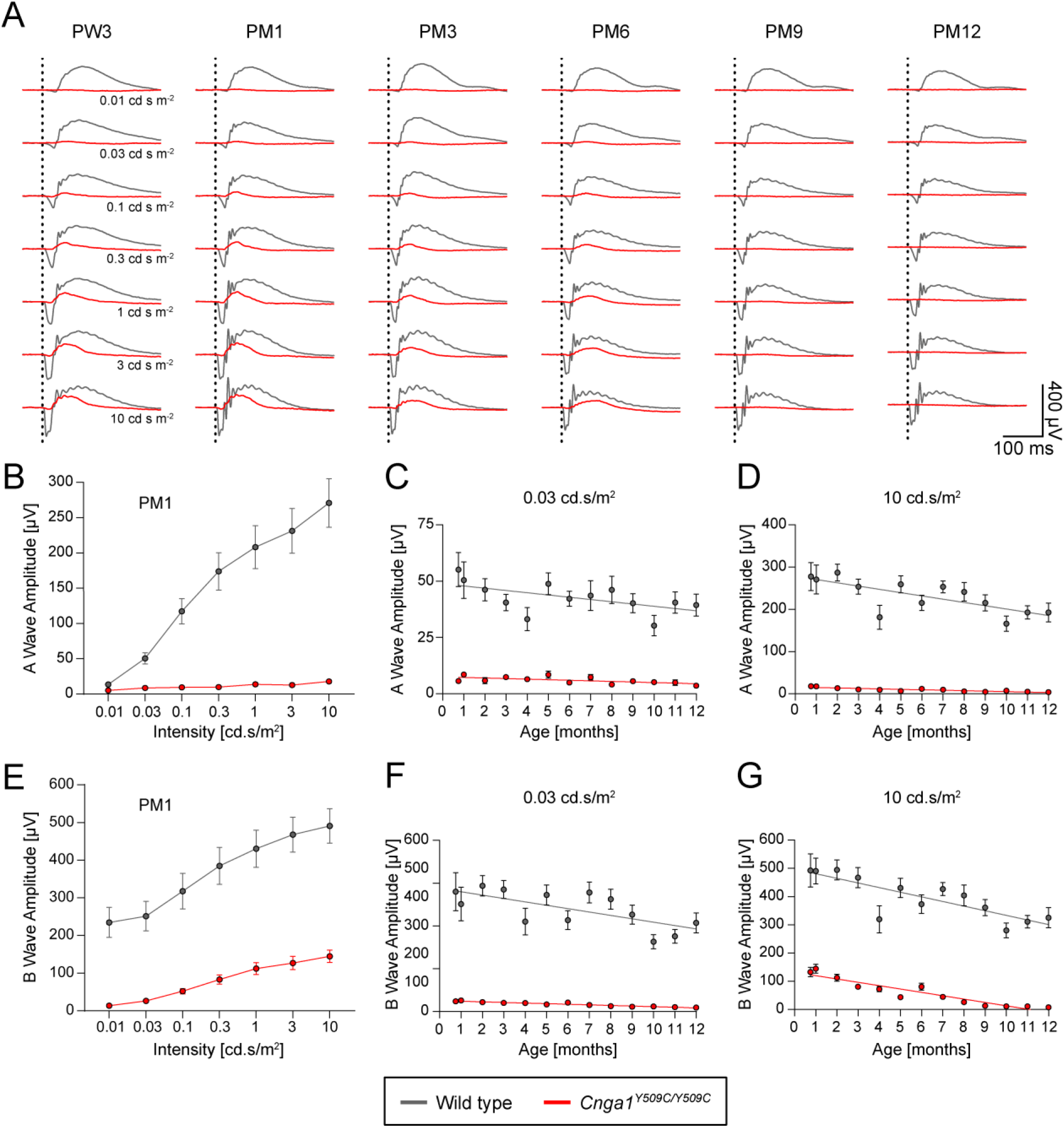
*Cnga1*^*Y509C/Y509C*^ mice show loss of rod-driven electroretinography. (**A**) Overlays of averaged ERG signals of *Cnga1*^*Y509C/Y509C*^ mice (red) compared to wild-type mice (black) at postnatal week 3 (PW3), postnatal month 1 (PM1), PM3, PM6, PM9 and PM12 at different light intensities. Vertical dotted lines mark the time points of light stimulation. Homozygous mutant mice show almost no (rod-mediated) response at low light intensities (0.01 and 0.03 cd s m-2) and also lose cone-mediated response between PM6 and PM9. (**B-G**) Quantification of a-wave and b-wave amplitudes of *Cnga1*^*Y509C/Y509C*^ mice (red) compared to wild-type mice (black) at different ages. a-wave (B) and b-wave (E) amplitudes were massively reduced in *Cnga1*^*Y509C/Y509C*^ mice compared to wild types for all light intensities at PM1. Rod-mediated responses for the a-wave (C) and b-wave (F) at a low light intensity (0.03 cd s m-2) as well as cone-mediated response for a-wave (F) at a high light intensity (10 cd s m-2) were reduced from the first measurement at PW3, while cone-mediated response for the b-wave (G) decreased over time. Wild-type mice: n=10; *Cnga1*^*Y509C/Y509C*^ mice: n = 12. Values are given as mean ± SEM.

### Rod degeneration in homozygous *Cnga1* mutant mice

The *Cnga1* mutation affects the morphology of the mouse retina (Fig. 6 A). While the overall lamination of the retina appeared to be preserved in the morphology of Cnga*1* mutant mice at PM1, PM2 and PM4, there was early shortening of the outer and inner segments of the photoreceptor layers, and at PM4, we found a marked thinning of the outer nuclear layer (ONL) (Fig. 6 A). Rhodopsin, a marker of rod outer segments, was stained in wild-type and *Cnga1*^*Y509C/Y509C*^ mouse retinas at PM1, PM3, PM6, PM9 and PM12. Indeed, the rhodopsin staining shows gradually reduction of rhodopsin expression in the mutant retina over time, revealing a compromised morphology of rod outer segments already at PM1 (Supplementary Figure 3).

**Fig. 6.**
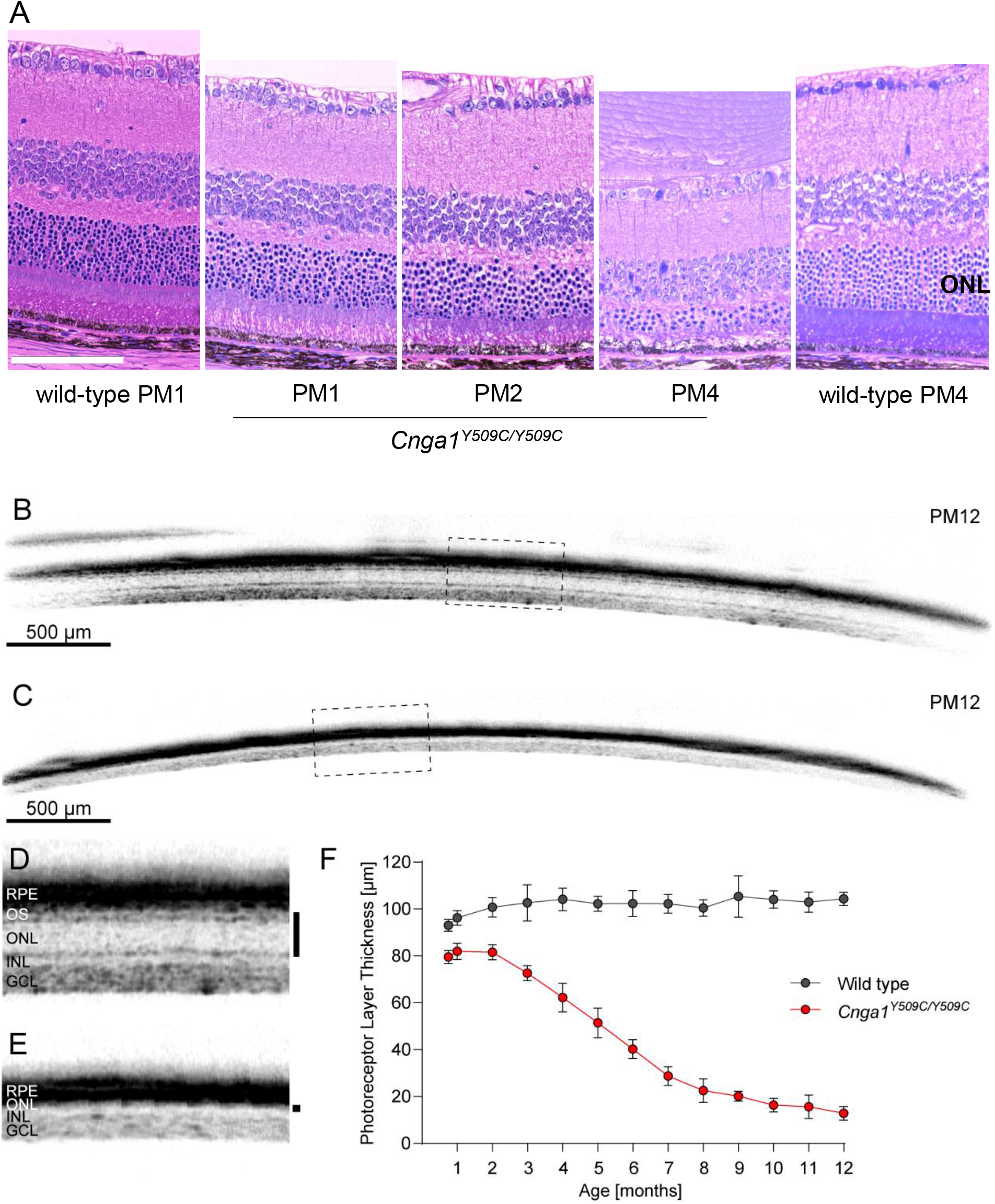
*Cnga1*^*Y509C/Y509C*^ mice show a reduced photoreceptor layer thickness. (**A**) Representative retina morphology images of wild-type mice and mutants showing progressive thinning of the ONL layer in the *Cnga1*^*Y509C/Y509C*^ mice. (**B-E**) Representative SD-OCT images of *Cnga1*^*Y509C/Y509C*^ and wild-type mice up to 12 months of age demonstrating a massive reduction of photoreceptor layer thickness in homozygous mutant mice (C, E) compared to wild-type mice (B, D). Black bars in close-ups D and E mark the thickness of the photoreceptor layer. RPE, retinal pigment epithelium; OS, outer segments; ONL, outer nuclear layer; INL, inner nuclear layer; GCL, ganglion cell layer. (**F**) Degeneration progress of photoreceptor layer thickness in homozygous mutant mice (red) compared to wild-type mice (black) from 3 weeks until 12 months of age. Wild-type mice: n=10; *Cnga1*^*Y509C/Y509C*^ mice: n = 12. Values are given as mean ± SD. Scale bar in A marks 100 µm.

To better characterize the progression of degeneration we performed a longitudinal *in vivo* spectral domain optical coherence tomography (SD-OCT) imaging study comparing *Cnga1* mutant mice to healthy age-matched wild-type mice (Fig. 6 B - F). SD-OCT evaluation of the entire photoreceptor length, measured as the combined thickness of the outer segment and the ONL layer (hereafter referred to as photoreceptor plus or PhR+), revealed an initial loss of 15 – 20 % of the PhR+ layer at PW3, PW4 and PM2 (Fig. 6 F). While the PhR+ layer thickness remained stable in the healthy control, it steadily declined over time in the *Cnga1* mutant (Fig. 6 F). Between PM5 and PM6 half of the PhR+ layer was lost in the mutant. Degeneration slowed after PM7, culminating in an almost complete loss of the PhR+ layer at PM12, and only the inner nuclear (INL) and ganglion cell (GCL) layers remained (Fig. 6 E, F). However, at PM12 the appearance of the INL nuclei starts to be more scattered, suggesting the appearance of secondary morphological changes in this inner retinal layer.

### Secondary degeneration of non-rod cells in *Cnga1* mice

*In vivo* BluePeak autofluorescence (BAF) and OCT angiography (OCT-A) were used to examine the morphology of the retinal fundus and the retinal vasculature (Fig. 7 A, B). At PM4, BAF imaging revealed accumulation of autofluorescent material in the fundus of *Cnga1* mutant mice (Fig. 7 A). In OCT-A scans, we observed altered vascular bed density in the mutant and a thinner appearance of the large blood vessels (Fig. 7 B). The SD-OCT and OCT-A measurements suggested the occurrence of secondary changes in non-rod cells as rod degeneration progresses. In order to analyse the morphology of cone photoreceptors over time, we labeled retinal sagittal sections of PM1, PM3, PM6, PM9 and PM12 wild-type and *Cnga1* mutant mice with the cone-specific marker peanut agglutinin (PNA), which labels the extracellular matrix of cone photoreceptors (20) (Fig. 7 C - H). At PM1, cone morphology, as judged by the PNA signal, appeared similar in both genotypes (Fig. 7 D). Over time, a gradual reduction of PNA signal was observed, revealing a loss of the outer segments of cones at PM9 and a complete loss of the cones at PM12 (Fig. 7 E – H).

**Fig. 7.**
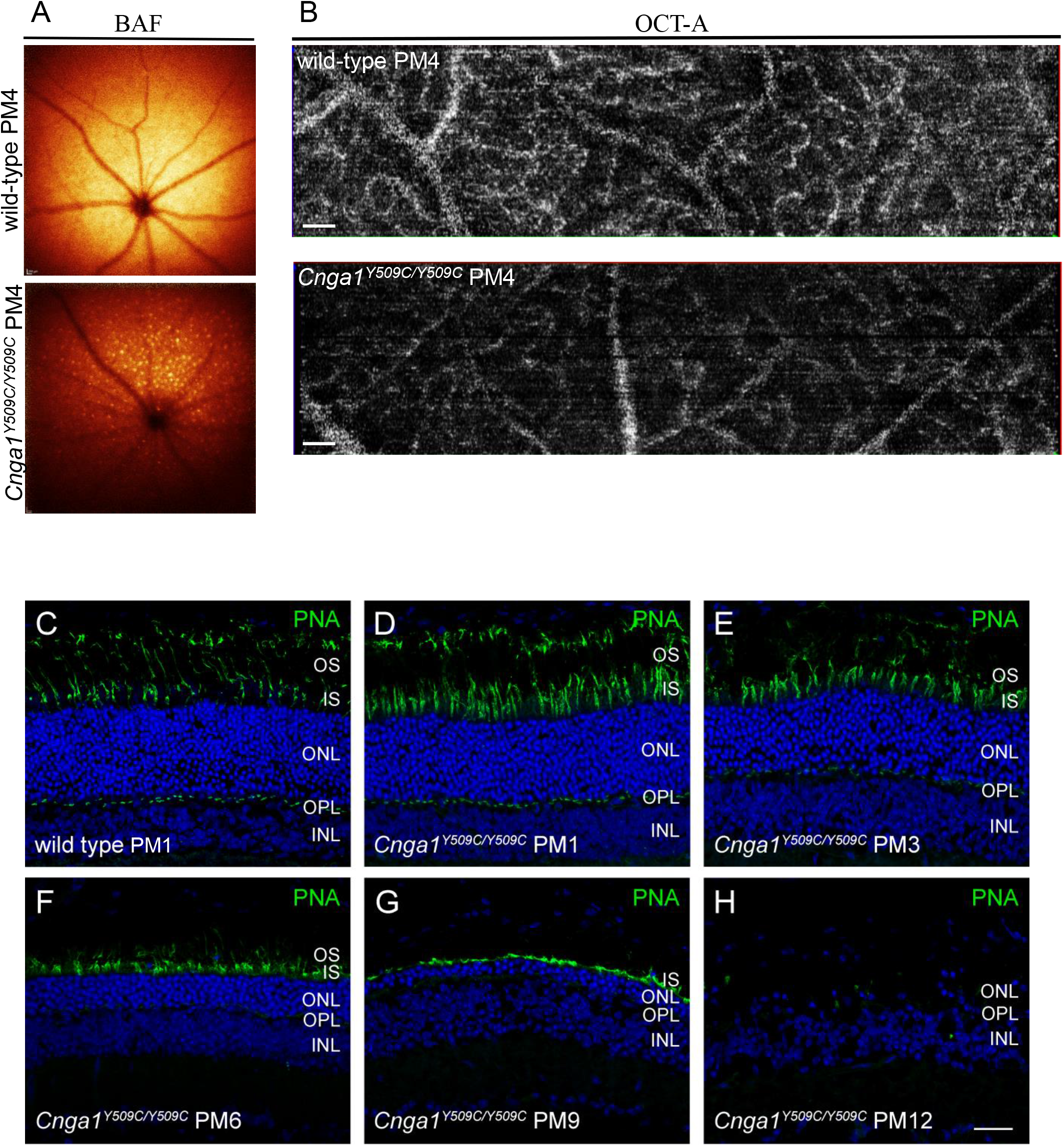
*Cnga1*^*Y509C/Y509C*^ mice show secondary retinal morphological changes and degeneration of cone photoreceptors. (**A-B**) Representative BAF and OCT-A scans of the fundus (A) and retinal vasculature (B) showing accumulation of autofluorescent spots and altered vascular bed in the *Cnga1*^Y509C/Y509C^ mouse fundus. (**C-H**) Representative confocal images showing expression of peanut agglutinin (PNA; green) in retinal cross sections of wild-type (1 month postnatal (PM1); (C) and *Cnga1*^*Y509C/Y509C*^ mouse retinas (D-H) at PM1, PM3, PM6, PM9 and PM12 demonstrating a degeneration of cone photoreceptors with age. Scale bars in B marks 200 µm. Scale bar in H marks 25 µm.

## DISCUSSION

In the present study, we have identified a family with autosomal recessive retinitis pigmentosa suffering from a homozygous c.1525G>A (NM_001379270.1) missense mutation in *CNGA1* leading to substitution of a Gly at position 509 by an Arg. So far, this mutation (previously recorded as c.1537 G > A; p.Gly513Arg (NM_000087.3)) has only been found as heterozygous in patients with retinitis pigmentosa (19). The same study described that transfection of HEK293 cells with the c.1537 G>A mutant *CNGA1* results in similar protein expression levels as the wild-type *CNGA1* (19). Gly509 is part of the loop linking two β-strands (β2 and β3) within the CNBD. While Gly509 is not directly involved in cGMP binding, it is most likely important for the flexibility of the β2/β3-connecting loop and thus for CNBD structure and function. In line with an important role, this glycine is conserved between species but also within the CNG channel family (4).

*CNGA1* was one of the first genes linked to RP over 27 years ago (21). To date, almost 50 probable pathogenic mutations in *CNGA1* have been identified (1) and a prevalence ranging of 2 % of arRP cases in Spain (22) to 7.6% in China (23) has been estimated. Despite its early discovery, little is known about *CNGA1*-RP, partly due to the lack of adequate animal models. More than 20 years ago a mouse model expressing an 890 bp *Cnga1* antisense DNA fragment was generated (24). A slight retinal degeneration was noted at 1 year of age (24). Since the effect of antisense expression was not confirmed at the protein level, it is unclear to what extent the observed morphological changes are due to a loss of CNGA1. Moreover, a functional characterization of this transgenic mouse line is missing. In 2015, the identification of a naturally occurring canine model with a mutation in the *Cnga1* gene was reported (25) providing a first genetic and clinical description. However, no detailed information on the retinal phenotype has been described. More recently, a mouse model with a targeted deletion in exon 2 of *Cnga1* was reported (26). This model carries an engineered 65 bp frame-shift deletion that, although not experimentally verified at the protein level, should result in a premature stop codon shortly after the deletion and loss of most of the ion channel protein. *Cnga1*-deficient mice were shown to lose the majority of photoreceptors by 16 weeks (26). Scotopic ERG responses to a single flash of 3 cd*s/m^2^ in these mice were greatly reduced at 3 weeks, which further decreased after 10 weeks (26).

Because the mutation identified in the here presented family results in a single amino acid exchange in the CNBD, this prompted us to develop an animal model that mimics this type of mutation. Indeed, we have identified from the ENU mutagenesis repository at the Helmholtz Center a mouse mutant with a c.1526 A>G mutation in *Cnga1*, resulting in a Tyr509Cys exchange in the CNBD of the CNGA1 protein. Tyr509 corresponds to Tyr513 in the human CNGA1 protein and participates in the formation of the β3 strand of the CNBD (4). This Tyr residue is conserved in the various mammalian CNG channel subunits and is even found in hyperpolarization-activated and cyclic nucleotide-gated channels (HCN), cGMP-regulated protein kinase 1 (PRKG1) or the cAMP-regulated protein kinase catalytic subunit (PRKACA). Interestingly, the structurally related potassium voltage-gated channel subfamily H member 1 (KCNH1), which contains a presumably non-functional CNBD and cannot be gated or modulated by cyclic nucleotides (27), has a Cys instead of Tyr at the corresponding position (aa 619). Although the mutation found in the ENU mouse model does not affect the same Gly residue as the c.1525 G > A mutation found in the patients, it affects a Tyr residue just four amino acids downstream from Gly509 and is thought to mimic the human Gly509Arg substitution.

Analysis of the CNG channel expression in 1 month old *Cnga1*^Y509C/Y509C^ mutant mice revealed an almost complete lack of CNGA1 protein, but normal *Cnga1* transcript levels. While *Cngb1* mRNA levels were also unaffected, we could not detect any CNGB1 protein in *Cnga1*^Y509C/Y509C^ at 1 month of age. This is in line with similar observations in *Cngb1*-deficient mice (16) and dogs (28), in which the absence of the CNGB1 subunit led to degradation of the remaining CNGA1 protein, although the *Cnga1* transcript was unaffected. It appears that the Tyr509Cys mutation has a major effect on protein structure and/or stability, resulting in a complete loss of the anti-CNGA1 immunosignal in Western blot and immunohistochemistry. Consistent with the loss of rod CNG channel function, we observed an absence of rod-driven ERG responses (a- and b-wave) from the earliest observation time point (PW3). This differs from what has been described in the *Cnga1* knockout mouse (26), but is most likely due to the fact that we used rod-specific ERG stimuli (e.g., the ISCEV rod stimulus: 0.03 cd.s/m^2^), while Liu et al. (26) used only a 3 cd.s/m^2^ stimulus, which elicits a mixed rod and cone response and therefore does not allow for segregation or distinct of rod-specific responses.

To gain a better understanding of the progression rate of rod and cone degeneration, we performed a 1-year *in vivo* imaging study with 13 observation time points. This allowed us to define with high granularity the time course of rod loss and the onset of secondary cone degeneration. Again, we observed differences between the *Cnga1*^Y509C/Y509C^ mouse line with a point mutation and the homozygous *Cnga1* knockout mouse. Liu et al. (26) showed retinal thickness measurements up to 4 months, while we followed the progression of retinal degeneration over 12 months. After 4 months, the thickness of the photoreceptor layer was reduced to only 20 % in the *Cnga1* knockout, but to about 60 % in the *Cnga1*^Y509C/Y509C^ mouse. The reason for the slow degeneration in these two mouse lines is not clear. In our Western blot and IHC analyses, we observed an almost complete loss of CNGA1 and CNGB1 proteins as early as 1 month. Since protein data are missing from the Liu et al. study (26), we can only assume that the rod CNG channel proteins are also missing in the *Cnga1* knockouts. In contrast to primary rod degeneration, secondary degeneration of cone photoreceptors appears to have a similar rate of progression in both mouse models.

In conclusion, single point mutations in the CNBD of the A subunit of the rod CNG channel are not tolerated and results in loss of both channel subunits at the protein level and eventual loss of function of this important ion channel of the rod photoreceptor transduction cascade. The *Cnga1*^Y509C/Y509C^ mouse appears to be an appropriate model of *CNGA1*-arRP and should be of great value for future studies on the molecular characterization of the pathobiology involved in the disease. Finally, the *Cnga1*^Y509C/Y509C^ mouse is a suitable model for preclinical proof-of-concept studies for future therapeutic approaches to treat this blinding disease.

## Data Availability

All data produced in the present study are available upon reasonable request to the authors.

## Acknowledgement

The authors thank Erika Bürkle, Monika Stadler and Andreas Mayer for expert technical assistance. SK thanks the German Academic Exchange Service (DAAD) (Funding no - 91525094) for her research stay in Germany (2014-2016), UGC -SRF fellowship grant and University of Madras. This work was supported by grants from the German Federal Ministry of Education and Research (Infrafrontier grant 01KX1012 to MHdA) and the German Center for Diabetes Research (DZD) (MHdA). The authors also thank the patient’s family towards their co-operation for this study.

## Data availability statement

All data relevant to the study are included in the article or uploaded as supplementary information. The raw data that support the findings of this study are also available from the corresponding author upon request.

## Figure legends

**Sup-Fig.1. Sequence chromatogram of affected family members:** *CNGA1* exon 10, depicting the homozygous mutation (c.1525G>A; p.Gly509Arg) in the affected family members (A, E, F), whereas all other unaffected family members (B, C, D, G, H) were heterozygous for this variant. The sequence of a control subject is given in I. The sites of the mutation are marked by arrows.

**Sup-Fig.2. *Cnga1*^*Y509C/Y509C*^ mice are lacking CNGA1 protein**: Western blot staining of *Cnga1*^*Y509C/Y509C*^ mouse retina at PM1 and PM6 using CNGA1- and CNGB1-antibodies. Β-Actin was used as control. N = 3 biological and technical replicates.

**Sup-Fig.3. *Cnga1*^*Y509C/Y509C*^ mice show compromised outer segments morphology**. (A-F) Representative confocal images showing expression of rhodopsin (green) in retinal cross sections of wild-type at PM1 (A), and *Cnga1*^*Y509C/Y509C*^ mouse retinas (B-F) at PM1, PM3, PM6, PM9 and PM12, illustrating the outer segment morphology. Rhodopsin staining shows a gradual reduction of rhodopsin expression in the mutant retina, revealing the compromised morphology of rod outer segments in the *Cnga1*^*Y509C/Y509C*^ retina already at PM1. Cell nuclei were stained with DAPI (blue). OS, outer segments; IS, inner segments; ONL, outer nuclear layer; OPL, outer plexiform layer; INL, inner nuclear layer.

