## Supplementary figures and images for "Mutations within the cGMP-binding domain of CNGA1 causing autosomal recessive retinitis pigmentosa in human and animal model"

### Supplementary Figure 2

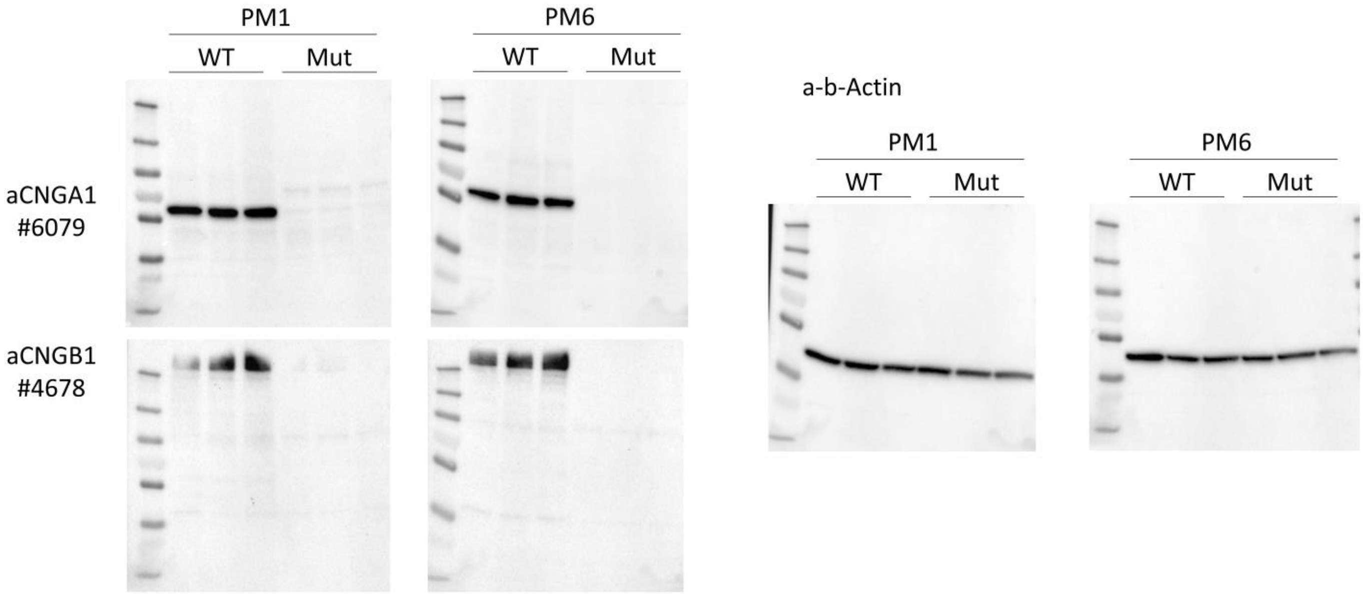

### Supplementary Figure 3

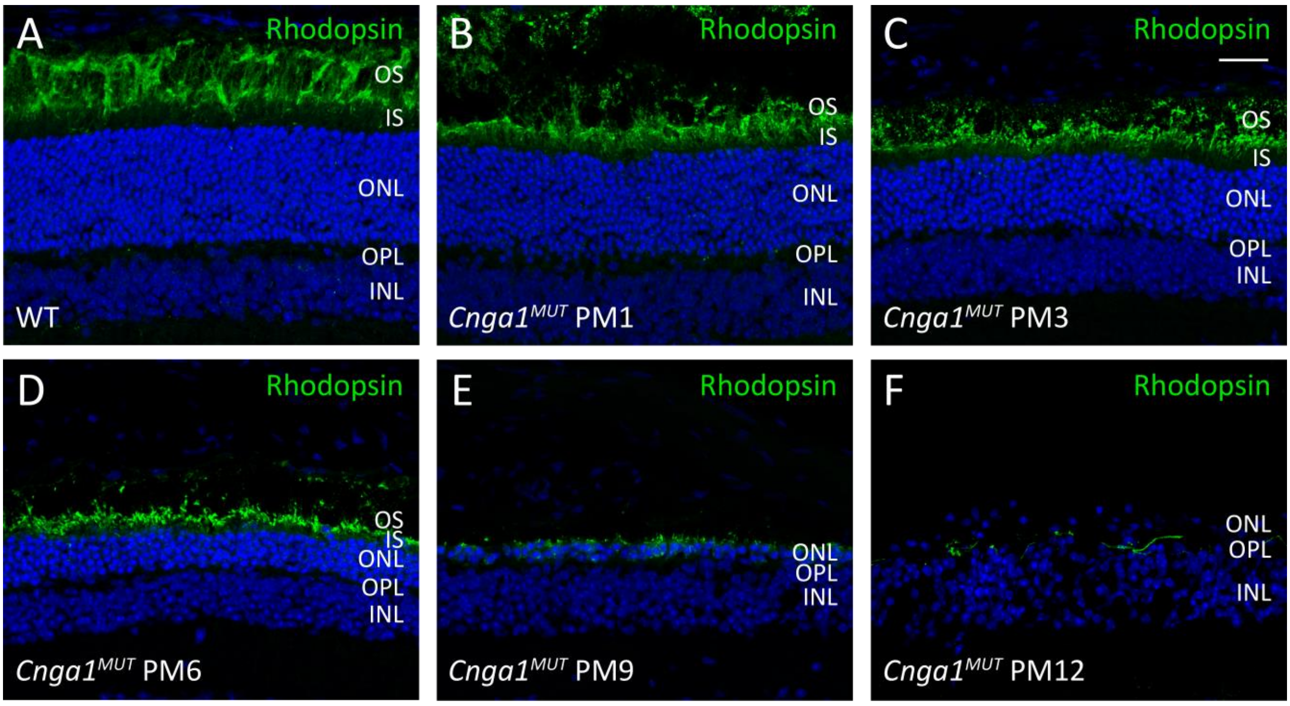

### Supplementary-Fig1

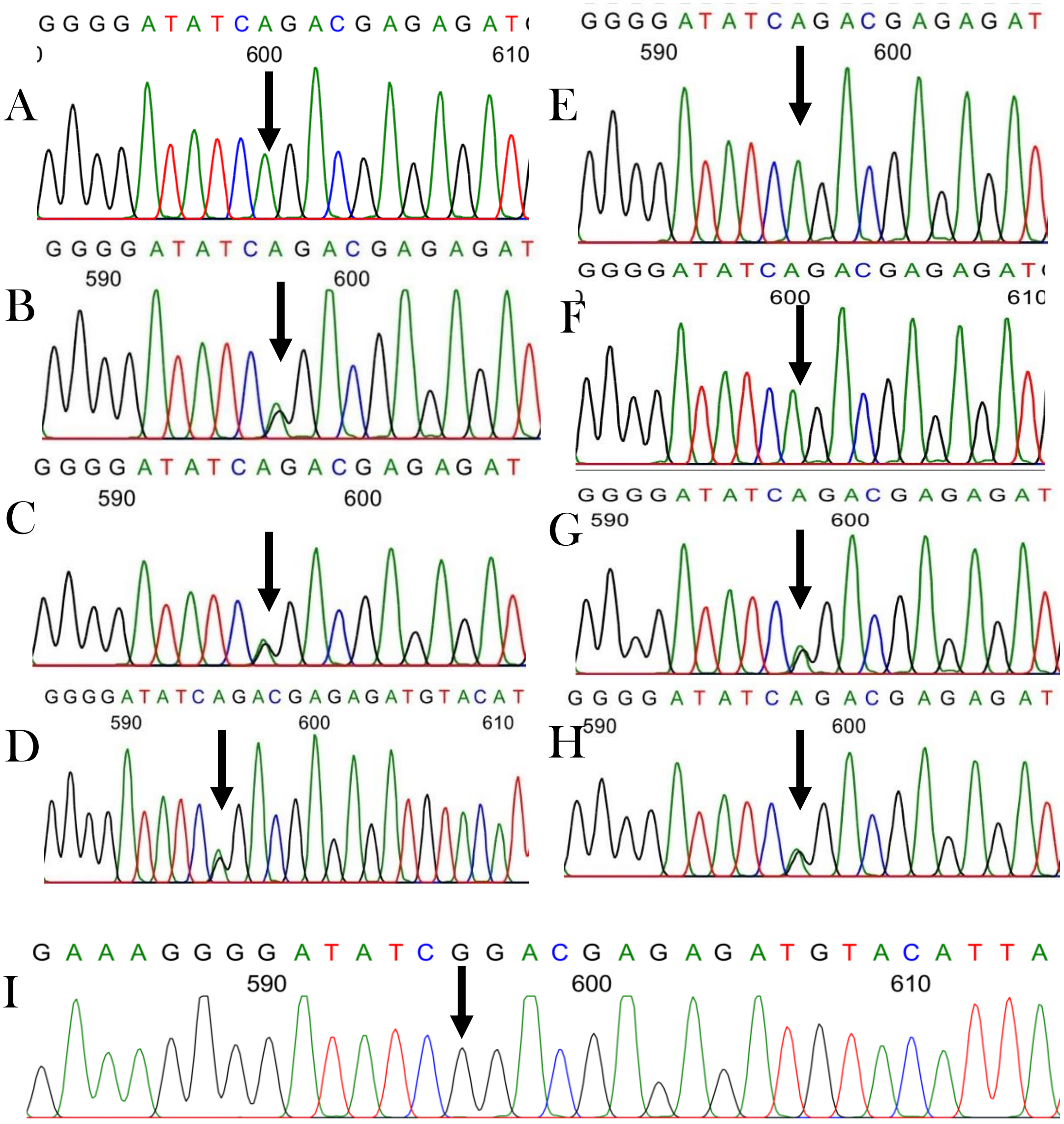
