## Supplementary table 1 for "Mutations within the cGMP-binding domain of CNGA1 causing autosomal recessive retinitis pigmentosa in human and animal model"

**Supplementary Tables of the Manuscript**

**Supplementary Table 1 - Retinal Degeneration Panel (105 Genes)**

| | | | | | | |
|---|---|---|---|---|---|---|
| *ABCA4* | *BEST1* | *DHDDS* | *IMPG2* | *OTX2* | *RBP3* | *SAG* |
| *ADAM9* | *C2ORF71* | *EYS* | *IQCB1* | *PDE6A* | *RD3* | *SEMA4A* |
| *AIPL1* | *CA4* | *FAM161A* | *KCNJ13* | *PDE6B* | *RDH12* | *SLC24A1* |
| *BBS1* | *CABP4* | *FSCN2* | *KCNV2* | *PDE6C* | *RDH5* | *SNRNP200* |
| *BBS10* | *CACNA2D4* | *GNAT1* | *KLHL7* | *PDE6G* | *RGR* | *SPATA7* |
| *TRIM32* | *CACNA1F* | *GNAT2* | *LCA5* | *PITPNM3* | *RHO* | *TOPORS* |
| *BBS12* | *CDHR1* | *GPR143* | *LRAT* | *PRCD* | *RIMS1* | *TRPM1* |
| *MKS1* | *CERKL* | *GPR179* | *LRIT3* | *PROM1* | *RLBP1* | *TTC8* |
| *CEP290* | *CLRN1* | *GRM6* | *MERTK* | *PRPF3* | *ROM1* | *TULP1* |
| *BBS2* | *CNGA1* | *GRK1* | *MKKS* | *PRPF31* | *RP1* | *UNC119* |
| *ARL6* | *CNGA3* | *GUCA1A* | *NMNAT1* | *PRPF6* | *RP2* | *USH2A* |
| *BBS4* | *CNGB1* | *GUCA1B* | *NR2E3* | *PRPF8* | *RP9* | *ZNF513* |
| *BBS5* | *CNGB3* | *GUCY2D* | *NRL* | *PRPH2* | *RPE65* | *EFEMP1* |
| *BBS7* | *CRB1* | *IDH3B* | *NYX* | *RAX2* | *RPGR* | *PITX2* |
| *BBS9* | *CRX* | *IMPDH1* | *OAT* | *RB1* | *RPGRIP1* | *FOXC1* |
