## Supplementary table 2 for "Mutations within the cGMP-binding domain of CNGA1 causing autosomal recessive retinitis pigmentosa in human and animal model"

**Supplementary Table 2 - Primers used in this study**

| **Primer** | **Sequence (5'- 3')** | **Ta** ($^0$C) | **Amplicon size (**bp**)** |
|---|---|---|---|
| *CNGA1-Ex-10-F* | GTATATCGTCATCATTATCCACTG | 60 | 855 |
| *CNGA1-Ex-10-R* | TCTGGGTACTCAGTTAGAGC | | |
| *Cnga1-Ex-9-F* | TGAGAGAGAAGTCCTGAGATACC | 63 | 400 |
| *Cnga1-Ex-9-R* | TGAGGTCATCTTTGGAGAGGC | | |
