## Supplementary table 3 for "Mutations within the cGMP-binding domain of CNGA1 causing autosomal recessive retinitis pigmentosa in human and animal model"

**Supplementary Table 3 – *CNGA1* gene variant identified in this study (HGVS nomenclature and variant classification applying ACMG Criteria)**

| **Chromosome** | **Position (hg38)** | **Ref Seq** | **Alt Seq** | **Gene** | **RS ID** | **HGVS Nomenclature** |
|---|---|---|---|---|---|---|
| chr4 | 47936957 | C | T | CNGA1 | rs544588016 | NM_001379270.1:c.1525G>A (p.Gly509Arg) |

| **ACMG Criteria** | **Justification** |
|---|---|
| PS3 Strong | Well established in vitro functional studies (Jin et al., 2016- PMID: 26802146). |
| PP3 Supporting | Pathogenic computational verdict based on 12 pathogenic predictions from BayesDel_addAF (Score - 0.4308), CADD (Score -26.1), DEOGEN2 (Score - 0.8299), EIGEN (Score -1.0079), FATHMM-MKL (Score -0.9805), LIST-S2 (Score -0.996), M-CAP (Score -0.7232), MVP (Score -0.9722), MutationAssessor (Score -4.1), MutationTaster (Score -1), PrimateAI (Score - 0.9237) and SIFT (Score -0) vs no benign predictions. – Meta predictors – All Meta predictors predicted this variant as pathogenic -REVEL (Score- 0.93); MetaLR (Score-0.9481); MetaSVM (Score-1.1063); MetaRNN (Score- 0.9325). |
| PM2 Moderate | Variant is present at extremely low frequency in the control population – gnomAD exomes - Allele frequency - 0.000008029 (2/249086) – No homozygotes – Present in only Asian population (East Asian – 1 in 17974; South Asian – 1 in 30588); gnomAD genomes (Allele frequency - 0.00001972 (3/152130)) {East Asians, 1 in 5194 alleles (0.0001925); European (Non Finnish, 1 in 68,040 (0.00001470); African/African American, 1 in 41,400 (0.00002415)} |
| PP1 Supporting | We have found this variant to co-segregate in autosomal recessive pattern (homozgyous) among individuals with RP in our family. Furthermore, the same variant reported to co-segregate in a Chinese family as compound heterozygous mutation along with another variation. |
| Classification | Pathogenic (PS3 Strong + PM2 Moderate + PP1 Supporting + PP3 Supporting) |
